# A first-in-human study in healthy subjects of the safety and pharmacokinetics of CNM-Au8, a suspension of catalytically active gold nanocrystals with remyelinating and neuroprotective properties

**DOI:** 10.1101/2023.05.11.23289871

**Authors:** KMS Kanhai, RGJA Zuiker, W Houghton, WG Kramer, K McBride, M Moerland, Adam Dorfman, Misty McGlothlin, M Hotchkin, M Mortenson, R Etherington, GJ Groeneveld

**Affiliations:** Centre for Human Drug Research, Leiden, the Netherlands; Clene Nanomedicine, Inc., Salt Lake City, United States; Clene Nanomedicine, Inc., North East, Maryland, United States; Kramer Consultancy LLC, North Potomac, Maryland, United States; Instat, San Diego, California, United States; Leiden University Medical Center, The Netherlands

**Keywords:** Gold nanoparticles, pharmacokinetics, remyelination, neurodegeneration, ALS

## Abstract

CNM-Au8 is a suspension of catalytically-active gold nanocrystals, developed to treat neurodegenerative disease. A first-in-human single- and multiple-ascending dose study was performed to assess safety and pharmacokinetics of CNM-Au8. The single-dose phase enrolled 40 subjects; the multiple-dose phase 48 subjects. Doses ranged from 15 to 90 mg CNM-Au8 and were administered by qualified clinical staff *per os*. Safety was assessed by adverse events, vitals, ECGs and laboratory measurements.

The most frequently reported related adverse event was mild abdominal pain (20%) of transient nature. Plasma half-life was 11.5 to 26.2 days, a first dose t_max_ ranged from 3.3 - 3.5 hours. CNM-Au8 displayed a safety profile and pharmacokinetic properties supportive of its advancement to Phase 2 and Phase 3 clinical trials.

## Introduction

Gold has been used in the treatment of various diseases since the beginning of civilization and is mentioned in documents dating as early as 2500 BC.^1^ Although it has been utilized for centuries, the exact mechanism of action of gold as a therapeutic agent remains uncertain.^1–3^

Currently, the gold complexes that are used as therapeutics in clinical practice (e.g., aurothiomalate and auranofin) are prescribed as immunomodulating therapies, predominantly in the treatment of rheumatoid arthritis.^4,5^ However, side effects including pruritus, dermatitis, stomatitis, diarrhea, proteinuria, and less frequently hematological abnormalities, are reported in up to 50% of patients and are a significant reason that individuals choose to discontinue therapy.^4,6^

Adverse events related to gold complexes may be specifically related to the covalent formulations of the gold complexes rather than to the activity of gold per se.^4,7^ Unlike aurothiomalate and auranofin, CNM-Au8 is not a gold salt. Instead, CNM-Au8^®^ is a highly concentrated suspension of non-covalently bound, clean-surfaced, faceted gold nanocrystals. The nanocrystals are produced as a result of a novel electrochemical crystallization method^8^, resulting in crystal lattices of pure gold atoms self-organized into various faceted, geometrical shapes (e.g., hexagonal bi-pyramid, pentagonal bi-pyramid, tetrahedron, and decahedrons).

CNM-Au8 gold nanocrystals have been shown to possess the catalytic activity of converting nicotinamide adenine dinucleotide hydride (NADH) into its oxidized form, NAD+.^9^ NADH and NAD+ constitute a key redox couple involved in energy-generating reactions in all cells, resulting in production of adenosine triphosphate (ATP). The NAD+ metabolic pathway has been identified as a novel therapeutic target for several neurodegenerative diseases.^10,11^ In mice treated with cuprizone, a well-characterized preclinical model for demyelination^12,13^, CNM-Au8 treatment enhanced remyelination and improved locomotor and gait behaviors.^9^ Based on the strength of this^9^ and other preclinical efficacy studies^14^ and a clean animal toxicity profile, CNM-Au8 is being developed for the treatment of demyelinating disorders such as multiple sclerosis (MS). It has also been shown to protect neuronal health and function by increasing ATP energy production and utilization, and is therefore also currently being investigated for the treatment of amyotrophic lateral sclerosis (ALS)^15^ and Parkinson’s disease (PD).

In this first-in-human study we aimed to assess the safety and pharmacokinetics (PK) of CNM-Au8.

## Methods

### Design

This was a first-in-human, randomized, double-blind, placebo-controlled study in healthy male and female subjects. The study had two phases: a single ascending dose (SAD) phase and a multiple ascending dose (MAD) phase. For each dose level in the SAD phase, 8 subjects were randomized in a 3:1 ratio to receive a single dose of either CNM-Au8 (n=6) or placebo (n=2). The initial dose level was 15 mg. Once interim safety evaluations were conducted and prespecified safety standards were passed prior to each dose escalation, additional cohorts of 8 subjects each were subsequently enrolled to investigate doses of CNM-Au8 at 30, 60, and 90 mg doses using the same 3:1 randomization scheme. Prior to the initiation of the second dosing cohort (30 mg) the Sponsor discovered that the initial 15mg batch had deviated from release specifications upon repeat testing and temporarily placed the study on hold. Following investigation of the deviation and subsequent production of replacement 15 mg study drug, the initial 15 mg SAD cohort was repeated with another 15 mg SAD cohort (n=6).

In the MAD Phase, approximately 48 subjects were planned for enrolment in 4 cohorts of 12 subjects each in a 3:1 ratio of active drug (n=9) to placebo (n=3) at the same dose levels of the SAD phase. Additional cohorts of 6 subjects (4 active and 2 placebo) for the SAD Phase and 9 subjects (6 active and 3 placebo) for the MAD Phase could have been added each time a dose needed to be repeated or adjusted downward according to the prespecified dose escalation rules. However, no additional cohorts were added based upon the dose escalation rules.

An interim safety analysis was conducted after completion of each cohort in both the SAD and MAD phase. Safety data, collected until one week after last dosing, was used in interim safety reports and was evaluated before it was decided to continue to the next cohort at the next higher dose level.

The study was approved by the Medical Ethics Committee of the BEBO Foundation (Assen, The Netherlands). The study was conducted according to the Dutch Act on Medical Research Involving Human Subjects (WMO) and in compliance with Good Clinical Practice (ICH-GCP) and the Declaration of Helsinki. This study was registered with ClinicalTrials.gov and was assigned the identifier NCT02755870.

### Subjects

Eighty-six healthy subjects between the age of 18 and 45 years (inclusive) were recruited to participate in this study via the research institute database and advertisements. All subjects gave written informed consent and were subsequently medically screened before entry into the study. A cotinine test was performed at screening to exclude smokers; participants were additionally not allowed to smoke and had to refrain from smoking throughout the study. Subjects were asked to not drink alcohol from 3 days before start of the study until the end-of-study follow-up visit. The use of medication was not allowed during the study period, with the exception of hormonal contraceptives. Subjects were required to use double-barrier contraception from two weeks before start of the study until 3 months after the study. Table S1 lists the full protocol full inclusion and exclusion criteria.

### Treatment

CNM-Au8 was manufactured and concentrated to 1 mg/mL (1,000 ppm) gold (Au) in 6.5 mM sodium bicarbonate (NaHCO_3_) buffered pharmaceutical grade water. The colour of the suspension was dark red to purple with a pH within the range of 8.3 to 9.3. Placebo consisted of 6.5 mM NaHCO_3_ in pharmaceutical grade water dyed with food colorants to match the red colour of active drug. The investigational product was administered orally in volumes of 15 mL, 30 mL, 60 mL, and 90 mL, for the 15 mg, 30 mg, 60 mg, and 90 mg doses, respectively.

### Dose selection

Using the Food and Drug Administration (FDA) guidelines^16^ a starting dose of 15 mg was selected. Multiple safety and toxicology studies across diverse animal species (with maximum feasible dosing studies of CNM-Au8 up to 90 mg/kg/day) did not demonstrate any acute or subchronic toxicities, and accordingly a maximum tolerated dose (MTD) was not identified. Possible toxicology signals from nonclinical animal studies were an incidental decrease of blood platelet count in minipigs, and accumulation of small amounts of gold in the kidneys without affecting renal function. During a 28-day study in dogs with 10 mg/kg/day, a half-life (T_1/2_) of 333 hours, a C_max_ of 11.9 ng/mL, and a T_max_ at 24 hours were observed. The canine toxicology findings resulted in a No Observed Adverse Effect Level (NOAEL) at 10 mg/kg/day. Based on the NOAEL, an equivalent human dose was determined to be approximately 5.4 mg/kg based on body-surface adjusted area. In a 65-kg human subject, the selected starting dose of 15 mg CNM-Au8 represented a safety margin of 20-fold. The CNM-Au8 dose levels used in this study were 15, 30, 60, and 90 mg CNM Au8 or matching placebo.

### Safety

A treatment emergent adverse event (TEAE) was defined as an adverse event (AE) that was observed after starting administration of the investigational treatment. If a subject experienced an event both prior to and after starting administration of the treatment, the event was considered a TEAE only if it worsened in severity (i.e., reported with a new start date) after starting administration of the specific treatment. All TEAEs were coded and classified by system organ class (SOC) and preferred term (PT) using the Medical Dictionary for Regulatory Activities (MedDRA version 17.0). All TEAEs collected during the investigational period were summarized by system organ class and severity.

Safety parameters included AEs, TEAEs, clinical laboratory assessments including urinalysis, vital signs, ECGs, and physical examinations. Concomitant medications were also described. Baseline was defined as the last non-missing value prior to dosing, except for ECG in which the baseline was defined as the average of the 3 baseline measurements. Change from baseline was calculated for all continuous safety parameters.

Twelve-lead ECG recordings were performed using Electrocardiograph Marquette 800/5500 or Dash 3000. Blood pressure and heart rate were assessed using a Nihon-Kohden BSM-1101K monitor or a Colin Pressmate BP 8800 or a Dash 4000. All ECG, blood pressure and heart rate measurements were performed after subjects had been resting in a supine position for at least 5 minutes.

In addition to standard haematological, chemical and coagulation laboratory measurements, exploratory kidney injury markers Kidney Injury Marker-1 (KIM-1), alpha-glutathione S-transferase (α-GST) and neutrophil gelatinase-associated lipocalin (NGAL)^17^ were measured in the multiple dose cohorts. For KIM-1 analysis, 2 × 2.0 mL of urine was collected in Sarstedt tubes per time point and stored at -80ºC. For α-GST 2 × 800 μL of urine was collected in Sarstedt tubes per time point, mixed with 200 μL urine stabilizing buffer (BIO85STB, Argutus Medical®) and stored at -80ºC. For NGAL analysis, 2 × 2.0 mL of urine was collected in Sarstedt tubes per time point and stored at -80ºC.

### Pharmacokinetics

Whole blood samples were taken for measurement of gold concentration in blood. In the SAD cohorts, blood for PK analysis was collected at 1, 2, 3, 4, 6, 8, 12, 24, 48, 72, 96,120, 144, 168, 192, 216, 240, 264, 288, and 312 hours after dosing (± 2 hours for blood samples collected from 24 hours onward). In the MAD cohorts, blood for PK analysis was collected during the day on days 1, 7, 14 and 21 (at 1, 2, 3, 4, 6, 8, 12 hours after dosing and 24 hours. During the other dosing days (days 3-6, 9-13, and 16-21) and on selected days after the last dosing (days 23-28, 32, 36, 40 and 49) one PK sample was taken per visit.

Blood was collected in tubes with potassium (K_2_-EDTA tubes), transferred to 2 mL Sarstedt tubes and stored at −80°C. Blood concentrations of CNM-Au8 were determined using a validated Inductively Coupled Plasma – Mass Spectrometer (ICP-MS) analytical method by Clene Nanomedicine (North East, Maryland, United States). The Lower Limit of Quantitation (LLOQ) used was 1.5 ng/mL for the SAD part and subsequently upgraded to 0.75 ng/mL for the MAD portion of the study after introduction of new ICP-MS instrument with greater sensitivity.

### Statistics

All PK parameters were calculated using non-compartmental analysis. Whole blood concentrations below LLOQ were taken as 0 for the calculation of the descriptive statistics for whole blood gold concentrations at each sampling time. All PK calculations were done and individual subject whole blood concentration-time graphs were prepared using SAS^®^ for Windows^®^ Version 9.4.

Individual PK parameters for CNM-Au8 were summarized with descriptive statistics. PK parameters such as the maximum observed plasma concentration (C_max_), time to C_max_ (T_max_), the area under the plasma concentration versus time from time 0 to time of measurable plasma concentration after 24 hours (AUC_0-24h_) and terminal-phase half-life (t½) were calculated using non-compartmental analyses and pharmacokinetic modelling.

## Results

The study was performed from April 2015 to October 2016. Cohort 1 (single dose of 15 mg) was repeated due to the shift to a new batch of CNM-Au8 during the study, as described in the Methods.

### Demographics

In both the SAD and MAD phases, cohorts were balanced by sex, meeting the protocol specification that no more than two-thirds of subjects should be one sex. The mean age across all 4 dosing cohorts ranged from 21.8 to 29.5 years in the SAD phase, and from 22.0 to 28.1 years in the MAD phase. Detailed baseline subject disposition is given in Table 1.

**Table 1.** Subject disposition per phase and dosage.

|  | Single dose cohorts |  |  |  |  | Multiple dose cohorts |  |  |  |  |
| --- | --- | --- | --- | --- | --- | --- | --- | --- | --- | --- |
|  | 15 mg | 30 mg | 60 mg | 90 mg | Placebo | 15 mg<sup>2</sup> | 30 mg | 60 mg | 90 mg | Placebo |
| N | 12<sup>1</sup> | 6 | 6 | 6 | 10 | 8 | 9 | 9 | 9 | 11 |
| Male (%) | 5 (41.7) | 3 (50.0) | 3 (50.0) | 2 (33.3) | 4 (40.0) | 5 (62.5) | 5 (55.6) | 4 (44.4) | 5 (55.6) | 7 (63.6) |
| Female (%) | 7 (58.3) | 3 (50.0) | 3 (50.0) | 4 (66.7) | 6 (60.0) | 3 (37.5) | 4 (44.4) | 5 (55.6) | 4 (44.4) | 4 (36.4) |
| Mean age (years) | 23.5 | 21.8 | 24.5 | 25.8 | 29.5 | 25.9 | 22.0 | 26.7 | 28.1 | 27.8 |
| Mean height (cm) | 176.4 | 173.7 | 179.3 | 169.3 | 173.3 | 174.2 | 181.2 | 174.0 | 174.2 | 177.8 |
| Mean weight (kg) | 69.8 | 66.8 | 77.2 | 64.7 | 75.6 | 70.0 | 76.2 | 70.8 | 72.5 | 77.3 |
| BMI (kg/m<sup>2</sup>) | 22.2 | 22.1 | 23.9 | 22.6 | 25.2 | 22.7 | 23.1 | 23.4 | 23.9 | 24.2 |

<sup>1</sup>Cohort 1a (n=6) was repeated as Cohort 1b (n=6) when ongoing stability tests indicated the first batch of administered CNM-Au8 failed to reach stability specifications. See text for details.
<sup>2</sup>Due to circumstances for cohort 5 (multiple dose, 15 mg) 10 subjects were enrolled (instead of the planned 12 subjects).

A total of 86 subjects were enrolled in the study, of which 83 were followed through to protocol completion. Three subjects in the MAD phase discontinued the study after dosing: one subject in the 15 mg MAD cohort retracted consent during the follow-up phase after completing dosing; one placebo subject in the MAD phase missed the last follow-up visit; and one subject in the 90 mg MAD cohort was withdrawn from the study after pregnancy was confirmed. The group of subjects that participated in this study was a healthy young adult population. Baseline subject disposition can be found in Table 1.

### Safety

For both the SAD and MAD phases of the study, routine clinical laboratory assessments, vital signs, ECGs, and physical examinations did not reveal clinically notable findings; none resulted in serious AEs or AEs leading to discontinuation of treatment. The most frequently reported TEAEs were in the classes of Nervous System Disorders and Gastrointestinal Disorders. The vast majority of TEAEs were Grade 1 severity (mild). There were no serious TEAEs, TEAEs leading to discontinuation of treatment, or TEAEs considered severe, life threatening, or resulting in death. Overall, no dose response relationship to TEAEs was observed in the SAD or MAD phase of the study; however, the frequency of headache and gastrointestinal TEAEs was higher in the 90 mg MAD treated subjects.

The treatment-related AEs reported by 3 or more subjects are summarized in Table 2. The most frequently reported AE for subjects who received CNM-Au8 was abdominal pain (20%) versus 10% for placebo-treated subjects. These symptoms were mild, mostly intermittent and all resolved without dosing change. For subjects who received placebo, the most frequently reported AE was headache (28%). Other frequently reported AEs were gastro-intestinal in nature: diarrhea (7 subjects with CNM-Au8, 1 subject with placebo), nausea (5 subjects with CNM-Au8, 1 subject with placebo) and discolored feces (3 subjects with CNM-Au8, 0 subjects with placebo).

**Table 2.** Summary of treatment-emergent adverse events reported by 3 subjects or more.

|  | Total<br>(Active:Placebo) | Single dose cohorts |  |  |  | Multiple dose cohorts |  |  |  | Placebo<br>(n=21) |
| --- | --- | --- | --- | --- | --- | --- | --- | --- | --- | --- |
|  |  | 15 mg<br>(n=12) | 30 mg<br>(n=6) | 60 mg<br>(n=6) | 90 mg<br>(n=6) | 15 mg<br>(n=8) | 30 mg<br>(n=9) | 60 mg<br>(n=9) | 90 mg<br>(n=9) |  |
| Subjects with any AE | 34:13 | 2<br>(16.6%) | 2<br>(33.3%) | 5<br>(83.3%) | 5<br>(83.3%) | 5<br>(62.5%) | 4<br>(44.4%) | 3<br>(33.3%) | 8<br>(88.9%) | 13<br>(61.9%) |
| Abdominal pain | 13:2 | 0 | 0 | 0 | 0 | 1<br>(12.5%) | 5 (55.6%) | 2<br>(22.2%) | 4<br>(44.4%) | 2<br>(9.5%) |
| Headache | 11:6 | 1<br>(8.4%) | 0 | 1<br>(16.7%) | 0 | 1<br>(12.5%) | 1<br>(11.1%) | 1<br>(11.1%) | 6<br>(66.7%) | 6<br>(28.6%) |
| Diarrhea | 7:1 | 0 | 0 | 3 (50%) | 0 | 1<br>(12.5%) | 1<br>(11.1%) | 0 | 3<br>(33.3%) | 1<br>(4.8%) |
| Nausea | 5:1 | 0 | 0 | 0 | 0 | 1<br>(12.5%) | 0 | 1<br>(11.1%) | 3<br>(33.3%) | 1<br>(4.8%) |
| Feces discolored | 3:0 | 0 | 0 | 0 | 0 | 0 | 0 | 0 | 3<br>(33.3%) | 0 |

The SAD 15 mg CNM-Au8 administered to Cohort 1a failed to meet product specifications with ongoing stability tests following product release. Therefore, a second SAD cohort (Cohort 1b) was recruited and dosing was repeated with a new batch of CNM-Au8 meeting release specifications. A comparison of TEAEs between Cohorts 1a and 1b was subsequently performed. TEAEs were reported by 3 (50.0%) out of 6 subjects in the initial 15 mg cohort (Cohort 1a) and 4 (66.7%) out of 6 subjects in the 15 mg replacement cohort (Cohort 1b). No subjects in the initial 15 mg cohort experienced a TEAE considered related to study drug by the Investigator, while 2 (33.3%) out of 6 subjects in the replacement 15 mg cohort experienced a TEAE considered related to study drug by the Investigator. The 2 TEAEs that were considered possibly related by the Investigator in Cohort 1b were headache and alopecia.

Because no reference ranges for normal values for KIM-1, α-GST, or NGAL have been established to date, observed values from this study were compared to data previously obtained from healthy volunteers in other studies (data on file). We did not identify any clinically relevant change from baseline of the three kidney injury markers (KIM-1, α-GST, or NGAL) across the four dosing groups of CNM-Au8 (15 mg, 30 mg, 60 mg, and 90 mg) over the course of the MAD Phase.

In summary, there were no laboratory or clinical findings, no serious AEs, and no AEs leading to discontinuation of treatment, and no AEs were considered severe.

### Pharmacokinetics

For the single dose cohorts, only one whole blood Au concentration was measured above the LLOQ (1.5 ng/mL), in one subject who received 15 mg CNM-Au8. Consequently, no PK analysis could be done for the SAD phase of the study, and the pharmacokinetic analysis was based completely on the MAD cohort data.

A concentration-time graph for gold plasma concentration at different dose levels of CNM-Au8 is shown in figure 1. Based on pre-specified fit criteria, the elimination t½ could be calculated for only 39% of the subjects and the geometric mean ranged from 277 to 628 hr (11.5 to 26.2 days). Steady-state plasma concentrations for all cohorts, based on the geometric mean whole blood concentrations, were reached by the end of the 2nd week of dosing (Day 14). The geometric mean whole blood concentrations from one week onward increased in a dose-related, but not dose-proportional, manner. This was also the case for days 14 and 21, where the increases in both C_max_ and AUC_(0-24)_ were less than dose-proportional, and the exponents for the power model for both parameters, ∼0.43, were considerably lower than 1, indicating a less than dose-proportional increase in exposure. Pharmacokinetic parameters per dose are shown in table 3.

**Table 3.** Overview of pharmacokinetic parameters per dose for multiple dose cohorts. *Geometric mean and geometric CV (%) except T_max_ for which the median and SD is reported. †Relative to the dose on the study day. ‡All concentrations were below LOQ and no PK parameters could be estimated.

|  | Parameter* | Multiple dose cohort<br>15 mg / day |  | Multiple dose cohort<br>30 mg / day |  | Multiple dose cohort<br>60 mg / day |  | Multiple dose cohort<br>90 mg / day |  |
| --- | --- | --- | --- | --- | --- | --- | --- | --- | --- |
|  |  | Mean | CV (%)<br>or SD* | Mean | CV (%)<br>or SD* | Mean | CV (%)<br>or SD* | Mean | CV (%) or<br>SD* |
| Day 1 | $C_{max}$ (ng/mL) | ‡ | ‡ | 2.818 | 113.94 | 1.853 | 105.57 | 5.678 | 53.11 |
| | $T_{max}$ (hr) † | ‡ | ‡ | 3.286 | 4.855 | 3.389 | 8.162 | 3.515 | 8.441 |
| | $AUC_{(0-24)}$ (hr x ng/mL) | ‡ | ‡ | 55.186 | 110.77 | 37.79 | 111.76 | 109.862 | 53.44 |
| Day 7 | $C_{max}$ (ng/mL) | 1.219 | 30.16 | 1.792 | 61.37 | 1.494 | 47.33 | 1.724 | 93.35 |
| | $T_{max}$ (hr) † | 4.864 | 8.551 | 5.007 | 8.858 | 3 | 4.183 | 6.283 | 7.567 |
| | $AUC_{(0-24)}$ (hr x ng/mL) | 20.335 | 50.33 | 30.524 | 146.5 | 19.32 | 352.38 | 14.48 | 610.6 |
| Day 14 | $C_{max}$ (ng/mL) | 1.399 | 32.96 | 1.641 | 65.94 | 1.975 | 74.27 | 2.27 | 75.81 |
| | $T_{max}$ (hr) † | 3.286 | 4.271 | 6.579 | 2.759 | 6.402 | 7.351 | 10.358 | 8.28 |
| | $AUC_{(0-24)}$ (hr x ng/mL) | 30.726 | 31.71 | 17.993 | 416.38 | 35.687 | 101.09 | 41.978 | 102.13 |
| Day 21 | $C_{max}$ (ng/mL) | 1.53 | 38.26 | 1.982 | 39.89 | 2.348 | 60.57 | 3.326 | 118.99 |
| | $T_{max}$ (hr) † | 12.126 | 8.526 | 10.964 | 17.505 | 14.469 | 15.189 | 7.619 | 7.386 |
| | $AUC_{(0-24)}$ (hr x ng/mL) | 32.317 | 44.76 | 41.41 | 40.69 | 50.267 | 62.42 | 66.017 | 120.7 |
| | $T_{1/2}$ (hr) | 276.694 | 35.47 | 512.941 | 61.55 | 355.75 | 29.89 | 628.487 | 17.84 |

**Figure 1.**
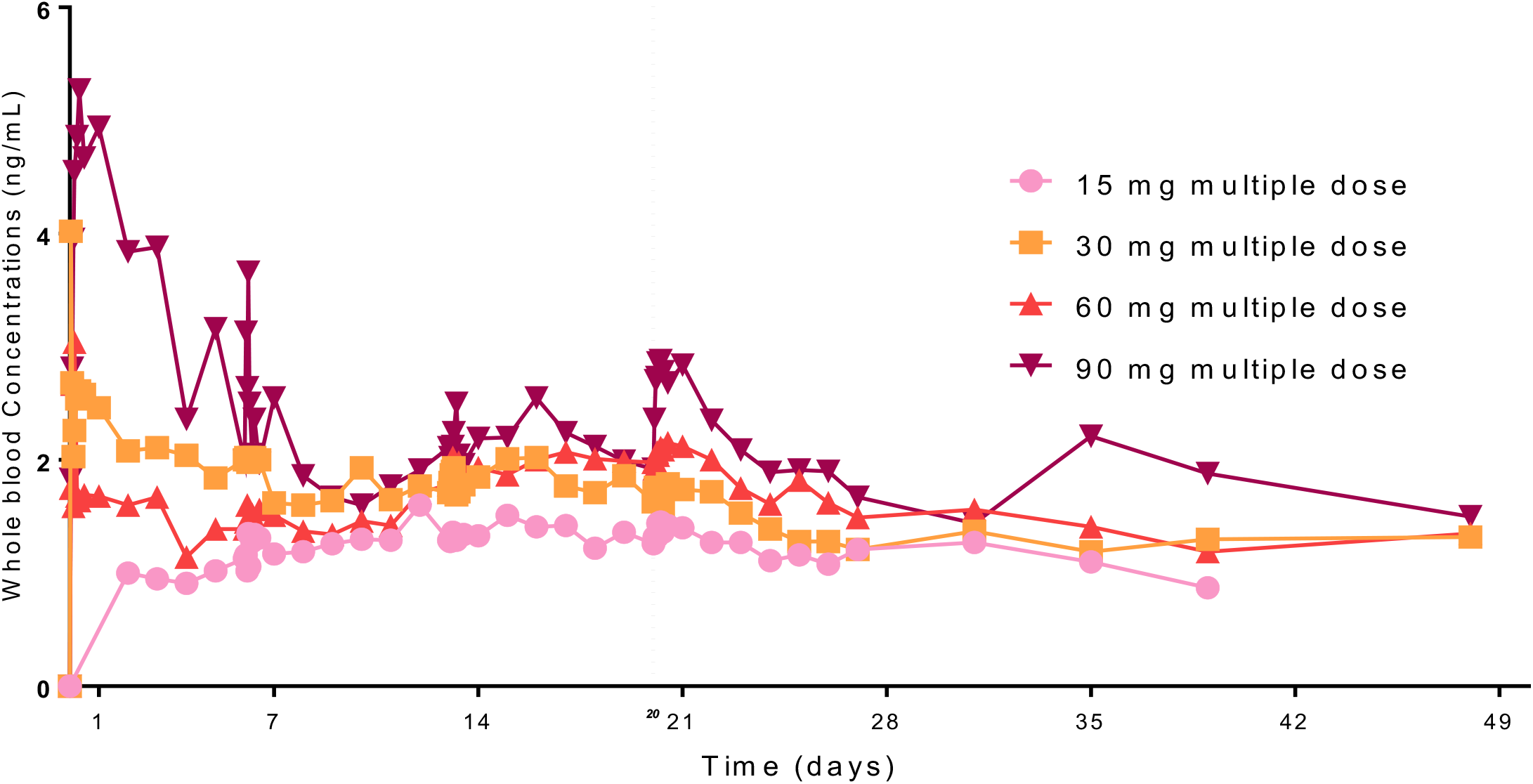
Geometric mean blood concentrations of gold after multiple oral administration of CNM-Au8 per dose level.

## Discussion

This first-in-human study assessed single (SAD) and repeated doses (MAD) of oral CNM-Au8 over 21 consecutive days. The studied doses ranged from 15 to 90 mg CNM-Au8; CNM-Au8 was considered safe and well tolerated. The main reported adverse event was mild self-limiting abdominal pain. Pharmacokinetics were characterised by a long half-life and increasing doses resulted in less than dose proportional exposure increases.

Gastro-intestinal (GI) adverse events (AEs) occurred in 32% of subjects after CNM-Au8 administration. This is comparable to the AE profile of auranofin, an oral gold-containing compound that leads to GI related AE’s in approximately 40% of cases.^1,18^ It is important to note that the treatment dosages of auranofin leading to adverse events are considerably lower than those of CNM-Au8; 3 – 9 mg compared to 15 - 90 mg respectively.^19^ This supports the hypothesis that the clean-surfaced, faceted gold nanocrystals comprising CNM-Au8, which has a uniquely different chemical composition than gold salts, is associated with fewer adverse events.^4,7^ GI side effects were rarely reported after intramuscular aurothiomalate administration leading to the assumption that route of administration may also be of importance.^1,20,21^

Another side effect often associated with gold salt-containing compounds is rash, which has been related with its parenteral administration. It has been reported in 12% subjects taking auranofin and in 30-50% for aurothiomalate.^1^ Although the exact mechanism is unknown, early studies report elevated IgE count in some patients^22^ and a significant relationship between rash as side effect and smoking.^23^ In our study, CNM-Au8 was orally administered and no rash was observed.

Literature reports 0-40% incidence of proteinuria due to use of auranofin or aurothiomalate^24^ and that the use of therapeutic dosages can affect renal tubular cells.^25^ This study did not detect any relevant increase or change from baseline across the 4 dosing groups of CNM-Au8 during the MAD phase for renal injury markers KIM-1, alpha-GST and NGAL. Because no abnormalities in urine related to kidney failure and no symptoms associated with renal insufficiency were observed, we conclude that no nephrotoxic effect of CNM-Au8 has occurred in this study. We further conclude that CNM-Au8 15 – 90 mg per day for 21 days was safe and well tolerated.

The low concentrations of CNM-Au8 measured in blood are in line with reported biokinetics of gold nanoparticles: gastro-intestinal absorption is limited (animal studies report incomplete absorption of less than 5% within 24h).^26,27^ The passage of gold nanoparticles across the gastro-intestinal epithelium appears to proceed in a prolonged fashion. There is an ongoing debate on the absorption route, but the most recent data suggest entry into the circulation might be through the lymphatic pathway.^27^ In animal studies, 10% of circulation nanoparticles is trapped by the liver after entry into the circulation and a part is redistributed again. The role of protein binding and interaction of circulating cellular elements on the biokinetics is not fully understood.^27^ Studies show a high accumulation of gold in secondary tissue, and gold detected 6 months after injection in expected organs (liver) and in less expected organs (brain, spinal cord, testis).^26–30^ This could explain the unusual PK curve of CNM-Au8 with gradually increasing concentrations, eventually reaching a plateau. Data suggest that biodistribution of gold nanoparticles is modulated over long periods of time, and it has been suggested that short-term pharmacokinetic studies should be interpreted with caution.^27^

CNM-Au8 is an aqueous suspension of gold nanocrystals, whose pharmacokinetic properties differ greatly from those of gold salts auranofin and aurothiomalate.^31^ It is clear that these registered compounds lead to higher blood concentrations of some form of gold, compared to that of CNM-Au8. Conversely, the long half-life that was reported for CNM-Au8 (11-26 days) is very comparable to that of auranofin (15-25 days), and of aurothiomalate (10-35 days).^1,32^ A long half-life can be explained by gold distributing widely into different tissues and it being chemically inert. The body cannot metabolise gold, and after distribution into the tissues it can be redistributed at a later time point.^33^

As CNM-Au8 was only orally administered without any intravenous dosing in the study, so absolute bioavailability and volume of distribution could not be determined. Therefore, these PK parameters of CNM-Au8 cannot be compared to other gold salt compounds such as the oral formulation of auranofin, which has a limited bio-availability of 20-30%.^31^

The size and shape of metal nanoparticles influence absorption in the intestine, accumulation, toxicity and interaction.^26,34,35^ The median nanocrystal ferret diameter of CNM-Au8 is 13 nm and can be expected to allow rapid absorption by intestinal epithelial cells compared to larger nanocrystal sizes, and has lower tendency to accumulate in these cells when compared to larger non-particles.^34^ The small size of the nanoparticle would suggest a good biodistribution throughout the body, compared to larger-sized particles (50-100 nm).

While none of the previous animal studies that were performed with CNM-Au8 reported a minimal anticipated biologic effect level (MABEL), we can estimate a human equivalent dose based on the AUC_0-24_ values in humans derived from the work reported here, the AUC_0-24_ values derived from the rodent chronic toxicology study, and the dosage used the demonstrated efficacy in two independent *in vivo* models of demyelination.^9^ These preclinical efficacy studies demonstrated that CNM-Au8 enhanced axonal remyelination and elicited functional/behavioral improvements in chemically-induced rodent models of demyelination. In these studies, rodents were dosed by gavage with CNM-Au8 at 10/mg/kg/day. Similarly, the dose of 10 mg/kg/day was investigated in a rodent 21-day toxicology study, in which it was shown that this dose resulted in an AUC_0-24_ exposure of 16.9 and 44.3 ng*hr/mL [mean males (n=6); females (n=6)]. Based on adjusted body-surface area, the 10 mg/kg CNM-Au8 by gavage per day dose corresponds to a human equivalent dose of 0.81 mg/kg/day.^36^ For a human subject weighing 65 kg this would be equivalent to approximately 53 mg/day, which was well in the range of dosages tested in this FIH study (15-90 mg per day). Further, based on the PK results from this study, the AUC exposure of AUC_0-24_ in humans at 21 days at 30mg/day of 41.4 ng*hr/mL can be compared to the average AUC_0-24_ 21-day rodent exposure at 10mg/kg of 30.6 ng*hr/mL. The human 21-day average AUC_0-24_ at the 30 mg/day dose represents approximately 1.35 times the 21-day animal exposure at 10 mg/kg/day, indicating that the 30mg human dose provides comparable exposure to CNM-Au8 to the exposure in rodents at the 10mg/kg/day dose that demonstrated treatment effects.

The use of injectable gold-salts has declined during the past decades.^1^ This is likely the result of the high occurrence of side effects in combination with the limited efficacy. In the treatment of rheumatoid arthritis (RA), methotrexate became the mainstay of treatment due to a superior benefit to risk ratio. In this clinical study the new gold nanocrystal formulation showed far fewer side effects than gold salts and demonstrated remyelinating and neuroprotective effects in nonclinical studies.^9^ The gold nanocrystals in CNM-Au8 were demonstrated to be an efficient catalyst for metabolic energy reactions, converting the energetic metabolite nicotinamide adenine dinucleotide hydride (NADH) into NAD+ in proof-of-principle cell-free assays.^9,14^ The increased availability of NAD+ and ATP, in addition to increased HSF1 activity, was shown to be neuroprotective in MS and amyotrophic lateral sclerosis (ALS) disease models.^37–41^ CNM-Au8 may offer an important possible treatment option for MS, Parkinson’s disease, and ALS.

This study did not include a pharmacodynamic analysis of the effects of CNM-Au8 on NAD+, NADH, or its ratio because these energy metabolite levels are tightly regulated under healthy conditions. Preclinical studies of the mechanism of CNM-Au8 demonstrated that the drug has little to no effect on myelination when healthy animals are treated with 10 mg/kg/day CNM-Au8.^9^ Furthermore, there is no effect on nervous system cell survival, function, or proliferation when healthy primary neuron or glial cells in culture are treated with CNM-Au8 with doses up to 10 μg/mL (final media concentration). In contrast, dose-dependent neuroprotective effects of CNM-Au8 are observed in culture starting at doses as little as 10 ng/mL when the neuron or glial cells are injured by neurotoxins, are derived from diseased animals, or are differentiated from induced pluripotent stem cells derived from patients.^14^ Because this study was performed in healthy subjects, we did not expect to see measurable changes in energy metabolites from biofluids; subsequent clinical studies of CNM-Au8 have included these biomarkers, such as plasma NAD+ and NADH, in their biomarker analysis plans and will be published when these data become available.

This study demonstrates that CNM-Au8 has a good safety profile, more favorable than old gold salt formulations. The next step in the development is to demonstrate CNM-Au8 possesses neuroprotective, neuroreparative effects *in vivo* and remyelinating effects as observed in nonclinical studies.

## Data Availability

All data produced in the present study are available upon reasonable request to the authors

## Acknowledgments

We are grateful the participants in this study for their generous donation of time and their commitment to contributing to clinical research. We thank Karen Ho, PhD, of Clene Nanomedicine for review of the manuscript. Clene Nanomedicine, Inc. sponsored this study.

## Summary points

- CNM-Au8 doses ranged from 15 to 90 mg were considered safe and well tolerated. The main reported adverse events were mild and self-limiting.
- The low concentrations of CNM-Au8 measured in blood are in line with reported biokinetics of gold nanoparticles: gastrointestinal absorption is limited and literature reports a high accumulation of gold in secondary tissue
- CNM-Au8 may offer an important possible treatment option for MS, and other neurological disease like Parkinson’s disease and ALS

## Tables

**Table S1.** Trial Inclusion and Exclusion Criteria.

|  |  |
| --- | --- |
| Inclusion Criteria | Female and male subjects were eligible for inclusion only if all the following criteria were met at screening: |
|  | 1. An IRB-approved informed consent was signed and dated prior to any study-related activities. |
|  | 2. Between the ages of 18 and 45 years, inclusive. |
|  | 3. Females were non-pregnant, non-lactating, and either post-menopausal for at least 1 year, surgically sterile (e.g., tubal ligation, hysterectomy) for at least 90 days, or agreed, from the time of signing the informed consent or 14 days prior to check-in until 90 days after Study Completion/Discharge, to use 2 forms of contraception. |
|  | 4. Body mass index (BMI) between 18 and 34 kg/m<sup>2</sup>. |
|  | 5. All laboratory values at screening were within normal range or were evaluated as not clinically significant (NCS) by the Investigator if outside normal range. |
|  | 6. Had no clinically significant abnormal findings during pre-dose physical examination or Screening and Baseline vital signs or ECG. |
|  | 7. Had not consumed and agreed to abstain from taking any dietary supplements or nonprescription drugs (except as authorized by the Investigator AND Medical Monitor) for 3 days prior to CRU admission through Follow-Up. |
|  | 8. Had not consumed and agreed to abstain from taking any prescription drugs (except as authorized by the Investigator AND Medical Monitor) during the 14 days prior to CRU admission through Follow-Up. |
|  | 9. Had not consumed alcohol-containing beverages starting 3 days prior to CRU admission and agreed not to consume alcohol through Follow-Up. |
|  | 10. Had not consumed grapefruit or grapefruit juice within the 14 days prior to CRU admission and agreed not to consume grapefruit or grapefruit juice through Follow-Up. |
|  | 11. Had not used tobacco- and nicotine-containing products within 60 days prior to the CRU admission and agreed to abstain from using tobacco- and nicotine-containing products through Follow-Up. |
|  | 12. Had the ability to understand the requirements of the study and was willing to comply with all study procedures. |
| Exclusion Criteria | A subject was **not** eligible for inclusion if any of the following criteria apply at screening: |
|  | 1. Had a history of illicit drug abuse in the past year or current evidence of such abuse in the opinion of the Investigator. |
|  | 2. Had positive findings on urine drug screen or cotinine test. |
|  | 3. Was positive for human immunodeficiency virus (HIV), hepatitis B and/or hepatitis C on screening assessments. |
|  | 4. Was pregnant or lactating. |
|  | 5. Had clinically significant medical or psychiatric history that, in the Investigator's judgment, would compromise the subject's safety or the collection of data. |
|  | 6. Had donated plasma within 7 days of CRU admission. |
|  | 7. Had lost or donated blood over 500 mL within 3 months (males) or 4 months (females) prior to screening. |
|  | 8. Had participated in a clinical trial within 30 days of screening or more than 4 times in the previous year (males) or more than 3 times a year (females). |

## References

1. Kean WF, Kean IRL. Clinical pharmacology of gold. Inflammopharmacology. 2008;16(3):112–125. doi:10.1007/s10787-007-0021-x

2. Stuhlmeier KM. The anti-rheumatic gold salt aurothiomalate suppresses interleukin-1beta-induced hyaluronan accumulation by blocking HAS1 transcription and by acting as a COX-2 transcriptional repressor. J Biol Chem. 2007;282(4):2250–2258. doi:10.1074/jbc.M605011200

3. Madeira JM, Bajwa E, Stuart MJ, Hashioka S, Klegeris A. Gold drug auranofin could reduce neuroinflammation by inhibiting microglia cytotoxic secretions and primed respiratory burst. J Neuroimmunol. 2014;276(1-2):71–79. doi:10.1016/j.jneuroim.2014.08.615

4. Dabrowiak JC. Metals in Medicine. Second edition. John Wiley & Sons, Inc; 2017.

5. Suarez-Almazor ME, Spooner CH, Belseck E, Shea B. Auranofin versus placebo in rheumatoid arthritis. Cochrane Database Syst Rev. 2000;2000(2):CD002048. doi:10.1002/14651858.CD002048

6. Menninger H, Herborn G, Sander O, Blechschmidt J, Rau R. A 36 month comparative trial of methotrexate and gold sodium thiomalate in the treatment of early active and erosive rheumatoid arthritis. Br J Rheumatol. 1998;37(10):1060–1068. doi:10.1093/rheumatology/37.10.1060

7. Yei Ho S, Tiekink ERT. 79Au Gold-Based Metallotherapeutics: Use and Potential. In: Gielen M, Tiekink ERT, eds. Metallotherapeutic Drugs and Metal-Based Diagnostic Agents. John Wiley & Sons, Ltd; 2005:507–527. Accessed August 22, 2014. http://doi.wiley.com/10.1002/0470864052.ch26

8. Mortenson MG, Pierce DK, Bryce DA, et al. Gold-Based Nanocrystals for Medical Treatments and Manufacturing Processes Therefor. Published online March 28, 2017.

9. Robinson AP, Zhang JZ, Titus HE, et al. Nanocatalytic activity of clean-surfaced, faceted nanocrystalline gold enhances remyelination in animal models of multiple sclerosis. Sci Rep. 2020;10(1):1936. doi:10.1038/s41598-020-58709-w

10. Lloret A, Beal MF. PGC-1α, Sirtuins and PARPs in Huntington’s Disease and Other Neurodegenerative Conditions: NAD+ to Rule Them All. Neurochem Res. Published online May 7, 2019. doi:10.1007/s11064-019-02809-1

11. Lautrup S, Sinclair DA, Mattson MP, Fang EF. NAD(+) in Brain Aging and Neurodegenerative Disorders. Cell Metab. 2019;30(4):630–655. doi:10.1016/j.cmet.2019.09.001

12. Torkildsen O, Brunborg LA, Myhr KM, Bø L. The cuprizone model for demyelination. Acta Neurol Scand, Suppl. 2008;188:72–76. doi:10.1111/j.1600-0404.2008.01036.x

13. Praet J, Guglielmetti C, Berneman Z, Van der Linden A, Ponsaerts P. Cellular and molecular neuropathology of the cuprizone mouse model: clinical relevance for multiple sclerosis. Neuroscience and biobehavioral reviews. 2014;47:485–505. doi:10.1016/j.neubiorev.2014.10.004

14. Wang Z, Henriques A, Rouvière L, et al. A Mechanism Underpinning the Bioenergetic Metabolism-Regulating Function of Gold Nanocatalysts. Neuroscience; 2023. doi:10.1101/2023.05.08.539856

15. Vucic S, Kiernan MC, Menon P, et al. Study protocol of RESCUE-ALS: A Phase 2, randomised, double-blind, placebo-controlled study in early symptomatic amyotrophic lateral sclerosis patients to assess bioenergetic catalysis with CNM-Au8 as a mechanism to slow disease progression. BMJ Open. 2021;11(1):e041479. doi:10.1136/bmjopen-2020-041479

16. U.S. Department of Health and Human Services Food and Drug Administration, Center for Drug Evaluation and Research. Guidance for Industry: Estimating the Maximum Safe Starting Dose in Initial Clinical Trials for Therapeutics in Adult Healthy Volunteers. Published online July 2005.

17. Bonventre JV, Vaidya VS, Schmouder R, Feig P, Dieterle F. Next-generation biomarkers for detecting kidney toxicity. Nat Biotechnol. 2010;28(5):436–440. doi:10.1038/nbt0510-436

18. Capparelli EV, Bricker-Ford R, Rogers MJ, McKerrow JH, Reed SL. Phase I Clinical Trial Results of Auranofin, a Novel Antiparasitic Agent. Antimicrob Agents Chemother. 2017;61(1):e01947–16. doi:10.1128/AAC.01947-16

19. Calin A, Saunders D, Bennett R, et al. Auranofin: 1 mg or 9 mg? The search for the appropriate dose. J Rheumatol Suppl. 1982;8:146–148.

20. van Riel PL, van de Putte LB, Gribnau FW, Macrae KD. Comparison of auranofin and aurothioglucose in the treatment of rheumatoid arthritis: a single blind study. Clin Rheumatol. 1984;3 Suppl 1:51–56. doi:10.1007/BF03342622

21. Roder C, Thomson MJ. Auranofin: repurposing an old drug for a golden new age. Drugs R D. 2015;15(1):13–20. doi:10.1007/s40268-015-0083-y

22. Iveson JM, Scott DG, Perera WD, Cunliffe WJ, Wright V. Immunofluorescence of the skin in gold rashes - with particular reference to IgE. Ann Rheum Dis. 1977;36(6):520–523. doi:10.1136/ard.36.6.520

23. Kay EA, Jayson MI. Risk factors that may influence development of side effects of gold sodium thiomalate. Scand J Rheumatol. 1987;16(4):241–245. doi:10.3109/03009748709102924

24. Kean WF, Hart L, Buchanan WW. Auranofin. Br J Rheumatol. 1997;36(5):560–572. doi:10.1093/rheumatology/36.5.560

25. Merle LJ, Reidenberg MM, Camacho MT, Jones BR, Drayer DE. Renal injury in patients with rheumatoid arthritis treated with gold. Clin Pharmacol Ther. 1980;28(2):216–222. doi:10.1038/clpt.1980.153

26. Schleh C, Semmler-Behnke M, Lipka J, et al. Size and surface charge of gold nanoparticles determine absorption across intestinal barriers and accumulation in secondary target organs after oral administration. Nanotoxicology. 2012;6(1):36–46. doi:10.3109/17435390.2011.552811

27. Schmid G, Kreyling WG, Simon U. Toxic effects and biodistribution of ultrasmall gold nanoparticles. Arch Toxicol. 2017;91(9):3011–3037. doi:10.1007/s00204-017-2016-8

28. De Jong WH, Hagens WI, Krystek P, Burger MC, Sips AJAM, Geertsma RE. Particle size-dependent organ distribution of gold nanoparticles after intravenous administration. Biomaterials. 2008;29(12):1912–1919. doi:10.1016/j.biomaterials.2007.12.037

29. Cho WS, Cho M, Jeong J, et al. Size-dependent tissue kinetics of PEG-coated gold nanoparticles. Toxicol Appl Pharmacol. 2010;245(1):116–123. doi:10.1016/j.taap.2010.02.013

30. Sonavane G, Tomoda K, Makino K. Biodistribution of colloidal gold nanoparticles after intravenous administration: Effect of particle size. Colloids and Surfaces B: Biointerfaces. 2008;66(2):274–280. doi:10.1016/j.colsurfb.2008.07.004

31. Blocka KL, Paulus HE, Furst DE. Clinical pharmacokinetics of oral and injectable gold compounds. Clin Pharmacokinet. 1986;11(2):133–143. doi:10.2165/00003088-198611020-00003

32. Furst DE, Dromgoole SH. Comparative pharmacokinetics of triethylphosphine gold (auranofin) and gold sodium thiomalate (GST). Clin Rheumatol. 1984;3 Suppl 1:17–24. doi:10.1007/BF03342618

33. Naz F, Koul V, Srivastava A, Gupta YK, Dinda AK. Biokinetics of ultrafine gold nanoparticles (AuNPs) relating to redistribution and urinary excretion: a long-term in vivo study. J Drug Target. 2016;24(8):720–729. doi:10.3109/1061186X.2016.1144758

34. Yao M, He L, McClements DJ, Xiao H. Uptake of Gold Nanoparticles by Intestinal Epithelial Cells: Impact of Particle Size on Their Absorption, Accumulation, and Toxicity. J Agric Food Chem. 2015;63(36):8044–8049. doi:10.1021/acs.jafc.5b03242

35. Carnovale C, Bryant G, Shukla R, Bansal V. Size, shape and surface chemistry of nanogold dictate its cellular interactions, uptake and toxicity. Progress in Materials Science. 2016;83:152–190.

36. Nair AB, Jacob S. A simple practice guide for dose conversion between animals and human. J Basic Clin Pharm. 2016;7(2):27–31. doi:10.4103/0976-0105.177703

37. Harlan BA, Pehar M, Sharma DR, Beeson G, Beeson CC, Vargas MR. Enhancing NAD+ Salvage Pathway Reverts the Toxicity of Primary Astrocytes Expressing Amyotrophic Lateral Sclerosis-linked Mutant Superoxide Dismutase 1 (SOD1). J Biol Chem. 2016;291(20):10836–10846. doi:10.1074/jbc.M115.698779

38. Nimmagadda VK, Makar TK, Chandrasekaran K, et al. SIRT1 and NAD+ precursors: Therapeutic targets in multiple sclerosis a review. J Neuroimmunol. 2017;304:29–34. doi:10.1016/j.jneuroim.2016.07.007

39. Penberthy WT. Nicotinic acid-mediated activation of both membrane and nuclear receptors towards therapeutic glucocorticoid mimetics for treating multiple sclerosis. PPAR Res. 2009;2009:853707. doi:10.1155/2009/853707

40. Homma S, Jin X, Wang G, et al. Demyelination, astrogliosis, and accumulation of ubiquitinated proteins, hallmarks of CNS disease in hsf1-deficient mice. J Neurosci. 2007;27(30):7974–7986. doi:10.1523/JNEUROSCI.0006-07.2007

41. Wang P, Wander CM, Yuan CX, Bereman MS, Cohen TJ. Acetylation-induced TDP-43 pathology is suppressed by an HSF1-dependent chaperone program. Nat Commun. 2017;8(1):82. doi:10.1038/s41467-017-00088-4

